# Factors affecting the quality of life of acne-suffering adolescents in Honduras, Central America, and its repercussions

**DOI:** 10.1101/2023.03.24.23287709

**Authors:** Eleonora Espinoza-Turcios, Kathya Chinchilla-Castañeda, Carlos Sosa-Mendoza, Lysien Ivania Zambrano, Henry Noel Castro Ramos, José Armada, Cristian R. Mejia

**Affiliations:** Institute for Research in Medical Sciences and Right to Health (ICIMEDES)/Scientific Research Unit (UIC), Faculty of Medical Sciences (FCM), National Autonomous University of Honduras (UNAH). Tegucigalpa, Honduras; Dermatology Service, Faculty of Medical Sciences (FCM), National Autonomous University of Honduras (UNAH). Tegucigalpa, Honduras; Honduran Institute for the Prevention of Alcoholism, Drug Addiction and Drug Dependency (IHADFA) Tegucigalpa, Honduras; Faculty of Business Sciences, Universidad Continental, Huancayo, Peru; Translational Medicine Research Centre. Universidad Norbert Wiener. Lima, Perú

**Keywords:** Acne Vulgaris, Adolescent, Central America, Quality of Life, Scar, Honduras

## Abstract

**Introduction:** Adolescence is a stage of physical, psychological, and social changes, which determine the personality of the individual, which can be influenced by physical appearance because of alterations in the skin texture of adolescents. Patients with dermatosis have a higher risk of developing depression, anxiety and suicidal ideas. Acne is an important factor for quality of life and affects adolescents both physically and psychosocially.

**Objective:** To determine the factors associated with the repercussions of acne in adolescents in Honduras.

**Methodology:** Analytical cross-sectional study. The Cardiff Acne Disability Index (CADI) scale was used through a questionnaire in Google Forms. Descriptive and analytical statistics were obtained.

**Results:** Of the 3,272 young participants, with respect to quality of life, 25.8% (845) had medium or high repercussions, 5.3% were very depressed by the appearance of their skin in the month prior to filling out the questionnaire. When multivariate analysis was performed, it was found that those who had medium or high repercussions due to acne were women (RPa: 1.33; 95%CI: 1.20-1.48; p-value<0.001), those of indigenous ethnicity (RPa: 1.36; 95%CI: 1.05-1.75; p-value=0.019), those who had scars on the face (RPa: 13.08; 95%CI: 10.02-17.05; p-value<0.001) or those who had the father (RPa: 1.13; 95%CI: 1.01-1.25; p-value=0.031) or siblings with acne (RPa: 1.16; 95%CI: 1.04-1.28; p-value=0.005).

**Conclusion:** It was found that gender, belonging to an indigenous ethnicity, having scars on the face or that having a close relative who had suffered from acne were associated with medium or high repercussions on the quality of life; it is important to take care of acne in this stage of life to avoid repercussions in adulthood.

## Introduction

Acne is a chronic inflammatory disease of the pilosebaceous unit, involving increased androgen-induced sebum production, abnormal keratinization of the pilosebaceous duct and an immune response to colonization by *Cutibacterium acnes* (formerly *Propionibacterium acnes*) (1, 2). Acne is estimated to affect 9.4% of the world’s population, making it the eighth most prevalent disease worldwide, affecting 90% of adolescents and young adults (3, 4).

It is known that adolescence is a crucial period for the development of social and emotional skills important for mental well-being, such as adopting healthy sleep patterns, exercising regularly, developing skills to maintain interpersonal relationships, coping with difficult situations, solving problems and learning to manage emotions. Therefore, it is important to have a favorable and protective environment in the family, school and community in general (5). The transition from childhood to adolescence entails physiological and psychological changes in the child (6), to which is now added the mental challenges generated by the COVID-19 pandemic, and it is important that adolescents receive the physical and mental care they need to develop, grow and thrive (7).

The visibility, chronicity and recurrent course of many skin lesions contribute to various psychosocial disorders, consequences of skin diseases. Patients with dermatoses are at increased risk of developing depression, anxiety, and suicidal ideation (4, 8, 9). In our society today, if certain social parameters are not met in physical appearance, adolescents suffer from bullying by their peers, acne can lead to emotional stress and the impact on quality of life can be significant (10).

There are studies that show the importance of acne, blemishes, scars and others that could have sequelae on the skin in adolescents (11), however, most of these studies have been conducted in Europe (12), the United States (13), India (14) and developed countries, in which a very important role is given to mental health and its influence on public health in general. On the other hand, in Latin American countries there are few studies that report this situation, as in Argentina and Mexico, but they were carried out in the pre-pandemic period (15, 16).

Among the few studies published in Honduras on the subject, in 2016 “Prevalence of dermatoses in school-aged children” was published, finding a prevalence of acne of 1.2% (17), to date no studies have been published in adolescent population related to acne. Therefore, the general objective is to determine the factors associated with the repercussions of acne in adolescents in Honduras.

## METHODOLOGY

The Scientific Research Unit (UIC) of the Faculty of Medical Sciences (FMC) - UNAH, implemented the methodology of national research on priority health problems in low-resource settings (18). The physicians who participated in the project were 65 eighth year medical students (MSS), who were trained in research methodology by the UIC-FCM, in dermatology with emphasis on acne and in quality of life by professors of the postgraduate courses of Dermatology and Psychiatry UNAH. After training, the surveyors were distributed in 16 departments of the country; this distribution was done by sampling in hospitals and primary care units, and information was collected from July to August 2020. Each interviewer provided an average of 58 questionnaires, each form had an average response time of 10 minutes.

To determine whether the number of respondents was sufficient to determine the association between the variables, the statistical power calculation was performed, where the only crosses that did not reach adequate power were the age of 13 years (power: 6%), for white ethnicity (power: 69%) and for whether the siblings had suffered from acne (power: 4%); therefore, only these 3 crosses should be taken with caution.

The study design was cross-sectional, analytical and multicenter, with a non-random snowball sample, which included 3325 young people aged 12-17 years; in the process of purging the database, 53 records were eliminated, 51 adolescents did not sign the informed consent form, and 2 records were not filled out in their entirety. The adolescents were children or family members within the staff of the care centers at where there were doctors doing. They were provided with the Google Forms link, and they socialized it with their peers. This type of survey was conducted due to the onset of the pandemic and considering the biosecurity measures, the restrictions of movement adopted by the country, the protocol, the safety of the surveyors and respondents. Therefore, it was adapted to online questionnaire and informed assent was obtained from each participant.

Before starting the project, ethical care was ensured, so the protocol was submitted to the FCM Biomedical Research Ethics Committee (CEIB), approval code: 010-2020, which was approved in the session of March 20, 2020. As part of their training, the MSS completed the online course on Good Clinical Practices The Global Health Network (www.tghn.org).

An instrument was prepared with the following sections: sociodemographic data including general data (age, sex, schooling, origin, ethnicity), as well as related factors and personal pathological history. To help determine whether they suffered from acne, two images were presented for the adolescent to identify their type of acne, inflammatory or non-inflammatory (comedogenic), and the Dermatological Questionnaire of Quality of Life in Adolescents (The Cardiff Acne Disability Index) was included (19).

Motley and Finlay described a brief questionnaire designed to assess disability caused by acne. The questionnaire adds a patient-oriented dimension to medical records, highlights patients with unusually high levels of disability, and increases the relevant information on which we base therapeutic decisions (20).

The Cardiff Acne Disability Index (CADI) is a questionnaire designed to measure the quality of life of adolescents and young adults with acne, we used the Spanish version validated in 2018, which was validated by Cardiff University. (19). As of October 2021, a change in wording was made (21).

When all the information was available, the database was cleaned and the variables were labeled and ready for analysis, which was performed with the statistical program Stata (version 16). First, we proceeded to generate three tables describing the population and their condition of acne, where we obtained the frequencies and percentages of the categorical variables, as well as the mean and interquartile range of age. Then the last tables were elaborated, where the RPc (crude prevalence ratios), RPa (adjusted prevalence ratios), 95% confidence intervals (95% confidence intervals) and p-values were obtained; all were obtained with the generalized linear models (Poisson family, log link function and models for robust variances). For a variable to pass from the crude model to the adjusted model, it had to have a p value <0.05; this same cut-off point was used to determine the final statistical significance.

## RESULTS

Of the 3272 respondents, 60.0% (1960) were female, the most frequent age was 16 years (25.1%), the median age was 15 years (interquartile range: 14-16 years), mestizos were the majority (74.4%) and they stated that their mother (20.2%), father (16.9%), brothers (37.7%), cousins (39.6%) and uncles (20.4%) had suffered from acne in their family.

When asked if they currently had acne, 52.5% (1717) responded that, if they did, of those who had acne, 44.3% (761) had it already more than 6 months ago, 19.7% (339) had inflammatory type acne and as for severity, 4.8% (82) and 25.7% (441) had severe and moderate acne, respectively. **Table 2**

**Table 1.** Characteristics of adolescents surveyed in Honduras, n=3272.

| Variable | Frequency | Percentage |
| --- | --- | --- |
| **Sex** |  |  |
| Male | 1306 | 40,0 |
| Female | 1960 | 60,0 |
| **Age (years old)** |  |  |
| 12 years | 190 | 5,8 |
| 13 years | 353 | 10,8 |
| 14 years | 454 | 13,9 |
| 15 years | 683 | 20,9 |
| 16 years | 820 | 25,1 |
| 17 years | 772 | 23,6 |
| Median and interquartile range | 15 years | 14-16 years |
| **Educational level** |  |  |
| Pre-basic or basic | 1415 | 45,7 |
| Intermediate level | 1681 | 54,3 |
| **Ethnic group** |  |  |
| Mestizos | 2391 | 74,4 |
| White | 342 | 10,6 |
| Other | 378 | 11,7 |
| Indigenous | 105 | 3,3 |
| **Family with acne\*** |  |  |
| Mother | 658 | 20,2 |
| Father | 553 | 16,9 |
| Brothers | 1229 | 37,7 |
| Cousins | 1293 | 39,6 |
| Uncles | 666 | 20,4 |
| **You have scars on your face** |  |  |
| No | 1701 | 52,0 |
| yes | 1571 | 48,0 |

\*This section does not add up to 100% since the questions were asked in a way that could have multiple answers.

**Table 2.** Current acne sufferers among adolescent respondents in Honduras: characteristics and impact, n=3272.

| Variable | Frequency | Percentage |
| --- | --- | --- |
| **Currently suffering from acne** |  |  |
| No | 1555 | 47,5 |
| Yes | 1717 | 52,5 |
| **Months suffering from acne\*** |  |  |
| < 1 month | 224 | 13,1 |
| 1-3 months | 452 | 26,3 |
| >3 months and up to 6 months | 280 | 16,3 |
| > 6 months | 761 | 44,3 |
| **It is inflammatory acne\*.** |  |  |
| No | 1378 | 80,3 |
| Yes | 339 | 19,7 |
| **Severity of acne\*** |  |  |
| Light | 1194 | 69,5 |
| Mild | 441 | 25,7 |
| Severe | 82 | 4,8 |

\*These questions have only been answered by those who mentioned that they had acne.

According to the impact test questions, 4.4% said that they felt very much aggressive, frustrated or embarrassed in the past month, and 3.4% thought that the acne they had in the past month interfered with their daily social life, social events or their relationship with people of the opposite sex. In addition, 5.3% were very depressed about the appearance of their skin in the past month and 25.8% (845) had medium or high repercussions from acne. Table 3

**Table 3.**
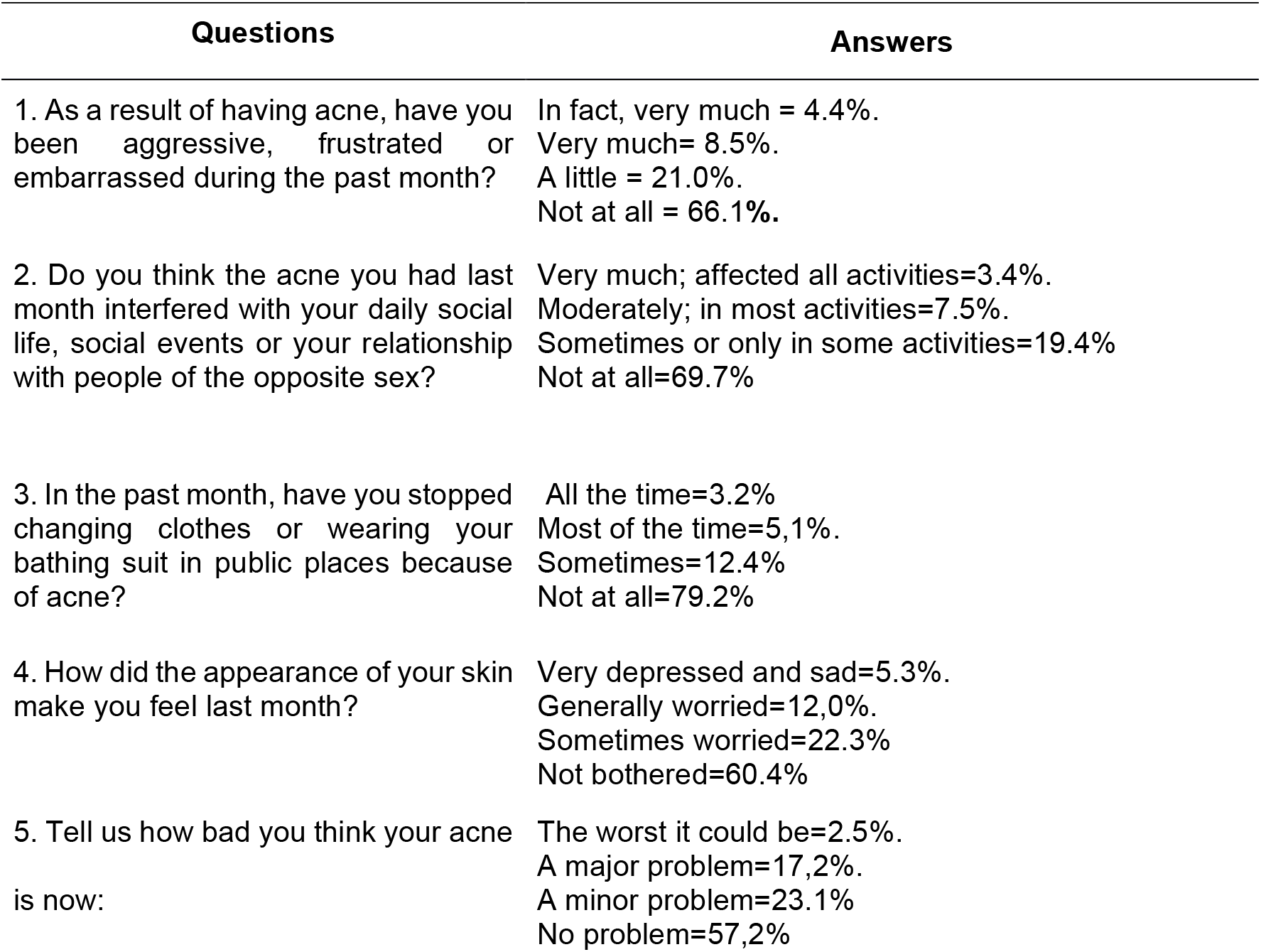
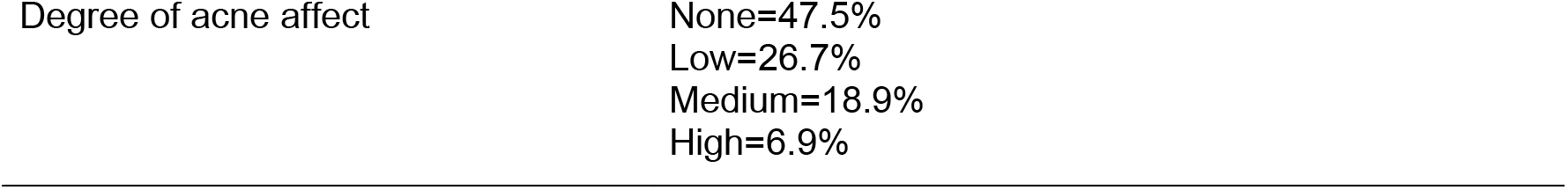
Disaggregated and global responses to the impact of acne among adolescent respondents in Honduras, n=3272.

| Questions | Answers |
| --- | --- |
| 1. As a result of having acne, have you been aggressive, frustrated or embarrassed during the past month? | In fact, very much = 4.4%.<br>Very much= 8.5%.<br>A little = 21.0%.<br>Not at all = 66.1%. |
| 2. Do you think the acne you had last month interfered with your daily social life, social events or your relationship with people of the opposite sex? | Very much; affected all activities=3.4%.<br>Moderately; in most activities=7.5%.<br>Sometimes or only in some activities=19.4%<br>Not at all=69.7% |
| 3. In the past month, have you stopped changing clothes or wearing your bathing suit in public places because of acne? | All the time=3.2%<br>Most of the time=5,1%.<br>Sometimes=12.4%<br>Not at all=79.2% |
| 4. How did the appearance of your skin make you feel last month? | Very depressed and sad=5.3%.<br>Generally worried=12,0%.<br>Sometimes worried=22.3%<br>Not bothered=60.4% |
| 5. Tell us how bad you think your acne is now: | The worst it could be=2.5%.<br>A major problem=17,2%.<br>A minor problem=23.1%<br>No problem=57,2% |

When bivariate analysis was performed, it was found that those who had medium or high repercussions due to acne were females (p<0.001), those who were 14 years old (p=0.022), 15 years old (p<0.001), 16 years old (p<0.001) or 17 years old (p<0.001), those of middle level (p<0, 001), those who had scars from acne (p<0.001) or those who had mother (p<0.001), father (p<0.001) or siblings with acne (p<0.001), on the contrary, those of other ethnicities had less impact from acne (p<0.001). **Table 4**

**Table 4.** Bivariate analysis of factors associated with the impact of acne in adolescents in Honduras, n=3272.

| Variable | Acne repercussions |  | Bivariate analysis |
| --- | --- | --- | --- |
|  | None or low | Medium or high | RPc (95%CI) p-value |
| **Gender** |  |  |  |
| Male | 1017 (77,9) | 289 (22,1) | Comparison category |
| Female | 1405 (71,7) | 555 (28,3) | 1,27 (1,13-1,45) <0,001 |
| **Age (years old)** |  |  |  |
| 12 years old | 165 (86,4) | 25 (13,2) | Comparison category |
| 13 years old | 305 (86,4) | 48 (13,6) | 1,03 (0,66-1,62) 0,886 |
| 14 years old | 358 (78,9) | 96 (21,1) | 1,61 (1,07-2,41) 0,022 |
| 15 years old | 468 (68,5) | 215 (31,5) | 2,39 (1,63-3,50) <0,001 |
| 16 years old | 582 (71,0) | 238 (29,0) | 2,21 (1,51-3,23) <0,001 |
| 17 years old | 549 (71,1) | 223 (28,9) | 2,20 (1,50-3,21) <0,001 |
| **Levels of education** |  |  |  |
| Pre-basic or basic | 1112 (78,6) | 303 (21,4) | Comparison category |
| Medium level | 1191 (70,9) | 490 (29,1) | 1,36 (1,20-1,54) <0,001 |
| **Ethnicity** |  |  |  |
| Mestizos | 1737 (72,7) | 654 (27,3) | Comparison category |
| White | 258 (75,4) | 84 (24,6) | 0,90 (0,74-1,09) 0,284 |
| Other | 310 (82,0) | 68 (18,0) | 0,66 (0,53-0,82) <0,001 |
| Indigenous | 72 (68,6) | 33 (31,4) | 1,15 (0,86-1,54) 0,348 |
| **Family with acne\*** |  |  |  |
| Mother | 409 (62,2) | 249 (37,8) | 1,65 (,147-1,87) <0,001 |
| Father | 323 (58,4) | 230 (41,6) | 1,83 (1,62-2,07) <0,001 |
| Brothers | 761 (61,9) | 468 (38,1) | 2,06 (1,83-2,31) <0,001 |
| Cousins | 935 (72,3) | 358 (27,7) | 1,12 (0,99-1,26) 0,057 |
| Uncles | 480 (72,1) | 186 (27,9) | 1,10 (0,96-1,26) 0,174 |
| **Scars on face** |  |  |  |
| No | 1644 (96,7) | 57 (3,3) | Comparison category |
| Yes | 783 (49,8) | 788 (50,2) | 14,97 (11,54-19,41) <0,001 |

\*Only the responses of those who, if they suffered, are shown; for the bivariate analysis, they are always compared with family members who did not suffer. RPc (crude prevalence ratios), 95% CI95% (95% confidence intervals) and p-values were obtained with generalized linear models (Poisson family, log link function and models for robust variances).

When performing the multivariate analysis, it was found that those who had medium or high repercussions due to acne were women (RPa: 1.33; 95%CI: 1.20-1.48; p-value<0.001), those of indigenous ethnicity (RPa: 1.36; 95%CI: 1.05-1.75; p-value=0.019), those who had scars on the face (RPa: 13.08; 95%CI: 10.02-17.05; p-value<0.001) or those who had the father (RPa: 1.13; 95%CI: 1.01-1.25; p-value=0.031) or siblings with acne (RPa: 1.16; 95%CI: 1.04-1.28; p-value=0.005). Table 5

**Table 5.** Multivariate analysis of factors associated with acne repercussions in adolescents in Honduras, n=3272.

| Variable | RPc (95%CI) p-value |
| --- | --- |
| **Gender** |  |
| Male | Comparison category |
| Female | 1,33 (1,20-1,48) <0,001 |
| **Age (years old)** |  |
| 12 years old | Comparison category |
| 13 years old | 0,83 (0,57-1,22) 0,345 |
| 14 years old | 1,05 (0,75-1,48) 0,768 |
| 15 years old | 1,37 (0,99-1,89) 0,055 |
| 16 years old | 1,25 (0,91-1,72) 0,171 |
| 17 years old | 1,25 (0,91-1,73) 0,170 |
| **Ethnicity** |  |
| Mestizos | Comparison category |
| White | 0,99 (0,84-1,17) 0,914 |
| Others | 0,86 (0,71-1,04) 0,112 |
| Indigenous | 1,36 (1,05-1,75) 0,019 |
| **Family with acne\*** |  |
| Mother | 1,05 (0,93-1,17) 0,419 |
| Father | 1,13 (1,01-1,25) 0,031 |
| Brothers | 1,16 (1,04-1,28) 0,005 |
| **Scars on face** |  |
| No | Comparison category |
| Yes | 13,08 (10,02-17,05) <0,001 |

The RPa (adjusted prevalence ratios), 95% confidence intervals (95% confidence intervals) and p-values were obtained with the generalized linear models (Poisson family, log link function and models for robust variances). Educational level was not included in the final model because it expressed the same as age, and variables that did not have a $p < 0.05$ in the bivariate model were not included.

## Discussion

The objective of the study was to determine the factors associated with the repercussions of acne in adolescents aged 12-17 years in Honduras, since there is no national data available. The only study on the subject found acne in eighth place among the most frequent dermatoses, with 1.2% in 59/15002 school-age children (17). A systematic review that included the opinions and experiences of people with acne vulgaris in 6 countries reported that the physical, psychological and social impact was common and often generated problems in forming new relationships and maintaining current ones (22).

Acne is an important factor for quality of life and affects adolescents both physically and psychosocially, they may have difficulties in establishing social relationships during this period, especially when so much importance is given to external appearance (23). In this study we found that one out of four young people perceived medium to high repercussions from acne on their face, knowing that skin conditions of varying severity can affect the quality of life of patients and have psychiatric consequences (24), Hazarika and Archana found facial acne as a single site involvement, this being the most common type (60%), but also with multi-site involvement (face, chest and back together) in 37% of cases, 75% had varying degrees of acne scarring, while post-acne hyperpigmentation was observed in 79% of cases (25). In Europe through an online survey on perceptions and management of acne, most people considered their acne (all stages of severity) to be either not a problem or a minor problem, although 29.7% considered it a major problem or burden (26).

We found in this study that females had more repercussions due to acne, a study in high school adolescents in Egypt, which included 994 students, found an overall prevalence of acne vulgaris of 33.5%, the mean age was 16.84±0.87, this being more common among females, with the mild form predominating, followed by moderate and severe with 6%. The CADI score was significantly related to disease grade and was highest among those with severe grade, followed by moderate and finally mild disease grade (27). Bosio et al. observed that the DLQI was higher in males than in females (15), contrasting our results with those found in this study, where women have a greater impact on quality of life due to acne. Therefore, this indicates that the suffering of this disease may vary according to the population, so studies should be done for each country or region.

Singh reported that “feelings of embarrassment and interference with social activities” scored significantly higher for women in a study of 1392 subjects with acne, using the Dermatologic Life Quality Index (DLQI) and the Cardiff Acne Disability Index (CADI) (28), the strongest association was facial scarring, which resulted in the highest frequency of acne-related repercussions, knowing that acne vulgaris can cause permanent physical scarring, negatively affect quality of life and self-image, and has been associated with higher rates of anxiety, depression and suicidal ideation (29). In Egypt, a study on the severity and impact on quality of life and self-esteem among adolescents according to the CADI score showed that 48.96% of students with clinically confirmed acne experienced a mild level of disability, while 11.46% of students with acne had a severe level of disability, low self-esteem was significantly more prevalent among females than males (30).

Acne vulgaris appears to share the same pathogenesis regardless of race or ethnicity but has different clinical presentations. In darker skin phototypes, there appears to be a greater subclinical inflammatory response, even in noninflammatory lesions, which is thought to trigger an exaggerated post inflammatory hyperpigmentation (PIH) response and keloid scarring (31), this study found that adolescents of indigenous ethnicity had fewer repercussions from acne. Among the adolescents studied aged 12-17 years, moreover, those who had a parent or siblings with acne also had more acne-related repercussions, knowing that positive family history is a strong predisposing factor in influencing acne presentation, severity and scarring (32). Parental acne was significantly associated with acne presentation and moderate/severe acne, while sibling acne was significantly associated with grade 3/4 scarring (33). A systematic review of the epidemiology of acne vulgaris showed a strong association between family history, age, BMI, and skin type and acne presentation or severity in multiple studies (34). A cross-sectional online survey study of individuals aged 15-24 years in Belgium, Czech and Slovak Republic, France, Italy, Poland, and Spain (n = 10,521) showed that a history of maternal or paternal acne was associated with an increased likelihood of acne (35). Among the risk factors correlated with an increased likelihood of scarring were acne severity, time between acne onset, first effective treatment, recurrent acne, and male sex (36). A study in a group of 15- to 35-year-olds reported that acne outbreaks located on the face were problematic for 81% of the study population (regardless of sex). Females experienced psychological distress more frequently than males (37). Jerome Kaikati et al. in a study conducted in Lebanon reported that acne creates a social barrier and is intensified in women of childbearing age (38). Therefore, people need support to understand the long-term management of acne, to develop control over acne and its treatments, to recognize the impact (22).

### Limitations

The collection of information was designed to be done in person in public middle schools throughout the country, however, due to the quarantine for the COVID-19 pandemic, it was modified and a Google Forms instrument was used, this through a non-random snowball sampling. It is in this context that the schools were forced to be at home, with restricted mobility and other characteristics of the pandemic. All this together could have had a bias at the time of recruitment and how they understood the questions, however, by using validated tests, the support of trained interviewers, the use of supporting images so that they could answer about the type of acne they suffered from and other ways that ensured a better approach to the respondents, all this together shows that we tried to minimize the bias to its minimum proportion. In addition, many adolescents were recruited, and although 3 crosstabs did not reach the necessary statistical power, only two of them had no association at the end of the analysis, so that the number of respondents was sufficient for the vast majority of the variable crosstabs that were raised.

## Conclusion

Based on the above, it is concluded that those who had a medium or high repercussion due to acne were women, those of indigenous ethnicity, those who had scars on their face or those who had a parent or siblings affected by acne.

## Data Availability

The data presented in this study are available on request from the corresponding author.

## Author Contributions

Conceptualization: EET, CASM and KCC

Data curation: CM, JA, EET, LIZ, and HNCR.

Formal analysis: CM, LIZ and JA.

Funding acquisition: KCC. Methodology: EET, CASM and KCC

Writing – original draft: EET, CASM, LIZ, KCC, CM, and JA.

Writing – review & editing: all authors have read and agreed to the published version of the manuscript.

## Institutional Review Board Statement

The study was conducted under the Declaration of Helsinki. This research’s preparation and execution fully complied with the fundamental ethical principles of autonomy, justice, beneficence, and non-maleficence. The Act Number (2020010), approved by the Ethics Committee in Biomedical Research (CEIB) of the National Autonomous University of Honduras (UNAH), meeting of March 20, 2020.

## Data Availability Statement

The data presented in this study are available on request from the corresponding author.

## Acknowledgments

All the MSS of the cohort October 2019 - October 2020 who accepted to participate in the project, to the directors of the healthcare centers in the areas that participated in the study, to the teachers of the UIC, for all the support provided in the review of the MSS assigned to each one. To Ovidio Padilla. who designed the online instrument and created the link for it, and Mauricio Gonzales for the data curation of the study.

